# User acceptability of saliva and gargle samples for identifying COVID-19 positive high-risk workers

**DOI:** 10.1101/2022.01.28.22270033

**Authors:** Kirsty McLennan, Ellen Barton, Christie Lang, Ian R. Adams, Gina McAllister, Martin A. M. Reijns, Kate Templeton, Ingólfur Johannessen, Alastair Leckie, Nick Gilbert

## Abstract

Throughout the COVID-19 pandemic nasopharyngeal or nose/throat swabs (NTS) have been the primary approach for collecting patient samples for the subsequent detection of viral RNA. However, this procedure, if undertaken correctly, can be unpleasant and therefore deters individuals from providing high quality samples. To overcome these limitations other modes of sample collection have been explored. In a cohort of frontline healthcare workers we have compared saliva and gargle samples to gold-standard NTS. 93% of individuals preferred providing saliva or gargle samples, with little sex-dependent variation. Viral titres collected in samples were analysed using standard methods and showed that gargle and saliva were similarly comparable for identifying COVID-19 positive individuals compared to NTS (92% sensitivity; 98% specificity). We suggest that gargle and saliva collection are viable alternatives to NTS swabs and may encourage testing to provide better disease diagnosis and population surveillance.

## Introduction

The World Health Organisation declared COVID-19 a global pandemic on March 11^th^ 2020 and called on all countries to ramp up their testing strategies. Unfortunately the COVID-19 virus remains a significant threat to public health as it continues to evolve, as has been seen for the emergence of the alpha (January, 2021), delta (June, 2021) and omicron (November, 2021) variants. Recent evidence suggests that omicron has reduced virulence compared to alpha and delta variants but omicron that has an increased transmission rate[1]. More variants are likely to arise, particularly in parts of the world that do not have good access to vaccines and large numbers of immunocompromised individuals. Testing therefore remains critical as part of a risk stratified approach to detect, isolate, and contain the virus, and will be key in facilitating the sustained reopening of society[2].

The recommended initial diagnostic sampling route for symptomatic individuals is combined NTS specimens tested using nucleic acid amplification tests (NAATs) such as quantitative reverse transcription PCR (RT-qPCR)[3]. However, in the UK and many other countries, individuals are recommended to use formal NTS testing in conjunction with lateral flow devices to facilitate rapid at-home testing. In PCR testing swab specimens are obtained from the nasopharynx and posterior pharynx/ tonsillar areas[4], whilst lateral flow devices use NTS or just nose swabs. Many find the procedure to collect NTSs uncomfortable or unpleasant which could impact uptake of, or compliance with testing and screening programmes. In particular this is likely to have a significant impact on asymptomatic testing. NTS sampling for PCR is also resource and labour intensive and testing capacity has been limiting in light of increased demand for tests and mass screening proposals. Furthermore, travel to a testing facility is often required to obtain a formal NTS and there is a risk of nosocomial transmission to the individual performing or facilitating the test due to the close contact required as well as the potential to induce involuntary coughing or sneezing. In order to overcome these barriers various alternative testing modalities have been explored.

Saliva has emerged as a promising alternative to nasopharyngeal swab testing as it is convenient, non-invasive, less resource intensive, and can be reliably self-administered. Saliva sampling is already an established practice in genetics to obtain nucleic acid samples, and has been used in the diagnosis of a number of respiratory viral infections prior to the COVID-19 pandemic, including other coronaviruses[5–7]. It has now been trialled in various healthcare settings internationally as an alternative diagnostic method in the detection of SARS-CoV-2[8–14]. Studies examining concordance rates of saliva with NTS testing have reported varying results – one study demonstrated increased sensitivity of saliva compared with NTS[15], while another reported that in a community setting saliva testing was less sensitive than NTS[16]. However, a recent meta-analysis of the available evidence concluded that saliva NAAT diagnostic accuracy is similar to that of NTS NAAT[17].

Pharyngeal gargle specimens have also been shown to be a useful sample type for detection of respiratory viruses including coronaviruses[7,18–20] and have shown comparability with NTS in the detection of SARS-CoV-2, although the available literature is more limited[21–23].

If practical to implement locally the use of saliva or gargle could be an alternative diagnostic modality for clinical staff and community testing, and be a means of increasing testing capacity and versatility. This mode of testing may also be well suited for the collection of samples from children, for example in a school setting, and for asymptomatic testing, for example those being routinely tested in the health and social care sector. We therefore set out to investigate the feasibility and utility of both saliva and pharyngeal gargle sampling methods, their relative acceptability, and their validity in the detection of the SARS-CoV-2 virus compared with nasopharyngeal testing. As samples are often stored before analysis we extended the study by exploring how sample storage conditions impacted test results.

## Results

### Detection of SARS-CoV-2 RNA in saliva and NTS samples

NTS testing is considered the gold-standard for SARS-CoV-2 diagnosis. However this sampling method is uncomfortable and deters individuals from regular testing. This is particularly challenging for high risk groups such as healthcare workers who are often exposed to patients with COVID-19 and who have to maintain a presence at work. Previously we developed a methodology to screen for SARS-CoV-2 in nasopharyngeal swabs stored in viral transport medium (VTM) collected from symptomatic individuals[24]. To determine whether saliva samples can be used for detecting SARS-CoV-2 RNA and to compare the specificity and sensitivity of NTS and saliva samples, the laboratory methodology was further adapted to facilitate viral RNA extraction from saliva (see methods). 109 healthcare workers provided NTS and saliva samples (Study Phase 1a) with an age range from 17 – 64 years (mean 40.2 (SD 1.2), median 41.0 (IQR 28.5-51.0). 79 were female (72.5%), and 29 were male (26.6%). Of the 109 paired samples there was a 0.9% (n=1) and 7.3% (n=8) amplification failure rate for NTS and saliva respectively, which may be due to high sample viscosity (Figure 1A). 10 NTS samples were found to be positive for SARS-CoV-2 RNA (Supplementary table 1), of these all-paired saliva samples were also identified as positive, whilst a further positive sample was identified, resulting in a total of 11 positive saliva specimens. This specimen had a relatively high Cq value in the TaqPath™ assays compared to other positive samples (33.5, 34.5, and 37.6 for the N, ORF and S genes respectively) (Supplementary table 1), however the distributions of Cq values for this small sample sets were similar (Figure 1B) indicating that saliva can be used for identifying COVID-19 positive individuals. Compared to NTS testing, sensitivity for saliva testing was 100% (95% CI, 69.1% - 100.0%) and specificity was observed to be 98.9% (95% CI, 94.0% - 99.97%).

**Figure 1.**
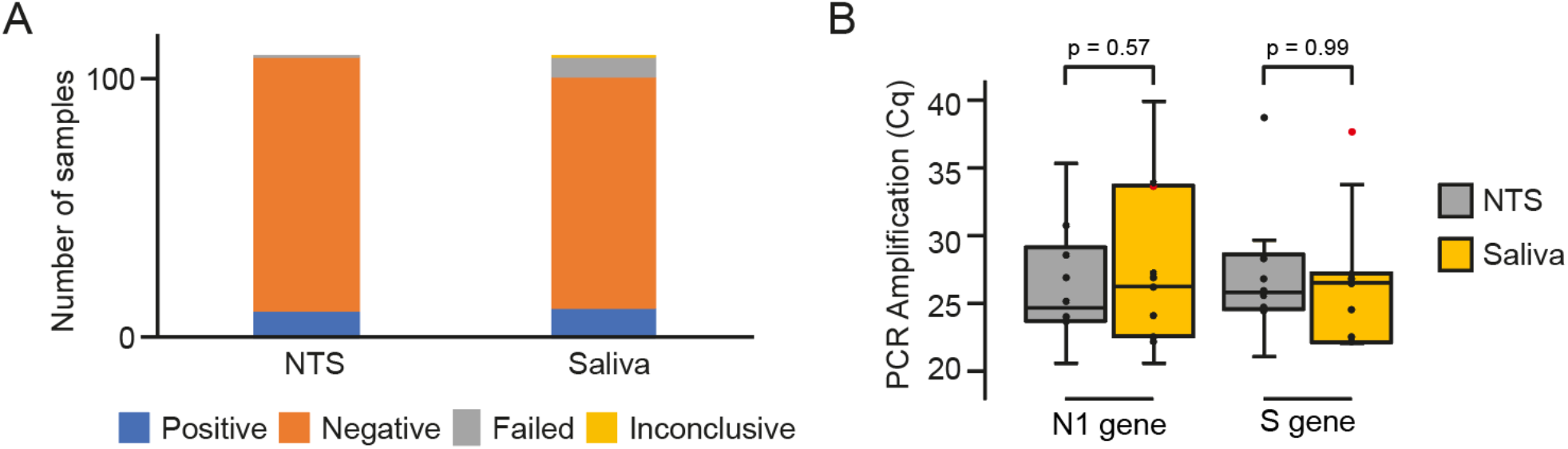
Identification of COVID-19 positive individuals using NTS or saliva samples. **(A)** Bar chart showing the proportion of positive, negative and failed tests in paired (n=109) NTS and saliva samples. **(B)** Boxplot showing the distribution of N1 gene and S gene Cq values for COVID-positive samples. Red point marks a sample identified as positive in the saliva sample but negative by NTS. P values are for a two tailed Wilcoxon test.

### Determination of optimal storage conditions for saliva samples

Often, patient samples are collected some distance from the testing lab necessitating samples to be stored for a period of time. To determine how storage conditions affected the ability to identify SARS-CoV-2 RNA in saliva samples a total of 206 participants each provided two saliva specimens and an NTS sample (Study Phase 1b). Saliva samples were then stored and transported at either ambient or cold (4°C) temperatures. The ages of the participants ranged from 6 – 66 years old (mean 37.7, SD 15.3; median 38.00,IQR 27-51), and included 147 (71.4%) females and 59 (28.6%) males. From these paired samples, 28 NTS specimens were found to be positive for SARS-CoV-2 (14%).

19 positive and 6 negative samples were selected at random and the cognate saliva samples shipped at ambient or cold temperatures were analysed and compared to the corresponding NTS results (Supplementary Table 2). Results were concordant between the two saliva samples stored under different conditions, but compared to the NTS samples only 17 samples were identified as being COVID-positive giving a sensitivity of 89.5% (95% CI, 66.9% - 98.7%) and a specificity of 100% (95% CI, 54.1% - 100.0%).

The objective of this phase was to compare how different shipping conditions might influence the ability of the laboratory to detect SARS-CoV-2 RNA, which presumably will be a reflection of viral RNA in the saliva samples. As noted, samples were concordantly called irrespective of shipping method, but it might be anticipated that due to RNA degradation at room temperature there would be a concomitant increase in Cq values. However, statistically there was no difference in Cq values between saliva samples stored at 4°C (Cold Chain) or at ambient temperature for the E gene (*P* = 0.57) or S gene target (*P* = 0.78) and the data distributions were similar (Figure 2).

**Figure 2.**
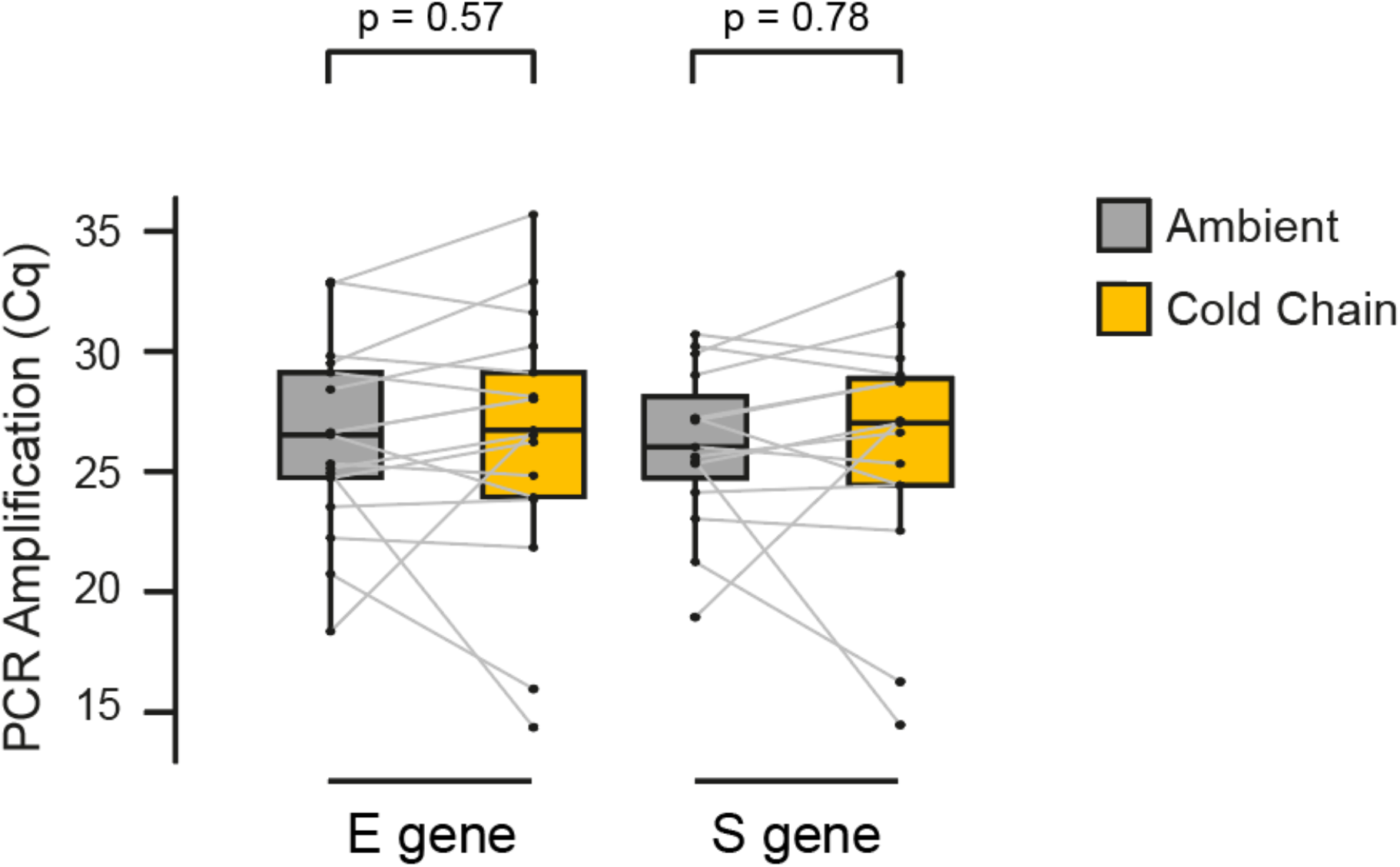
Effect of shipping conditions on SARS-CoV-2 RNA stability in saliva samples. Comparison of Cq values (E gene and S gene) in paired saliva/NTS samples following shipment to the laboratory under room temperature (RT) or cold chain (CC) conditions. P values are for a two tailed Wilcoxon test.

### User acceptability of saliva and gargle sample for SARS-CoV-2 RNA detection

During the COVID-19 pandemic frontline health workers have been regularly tested, predominantly by NTS. Although these individuals know the benefits of testing there is a risk that due to the unpleasant nature of taking nasopharyngeal swabs thoroughly, as well as testing fatigue, that over time adherence or sample quality might decrease. We therefore initiated a large testing programme (Study Phase 2) to explore alternative COVID-19 testing modalities by comparing the user acceptability and results of dependent NTS, gargle and saliva samples. Samples were collected from 261 individuals with ages ranging from 8 – 67 years old (mean 38, SD 14; median 37, IQR 22) and a gender breakdown of 22.2% male and 77.8% female.

Out of 261 individuals 46 (18%) were found to have NTS specimens positive for SARS-CoV-2 RNA. 37 positive and 30 negative NTS specimens were selected at random and the cognate saliva and gargle specimens were analysed by RT-qPCR. Internal control amplification failed in 3% of gargle samples and 9% of saliva samples, despite all being positive for the human RPP30 housekeeping gene^25^.

After discounting inhibited samples there were 65 NTS/gargle pairs (Supplementary data 3). 62 of the 65 NTS/ gargle pairs were concordant (34 positive and 28 negative pairs) whilst SARS-CoV-2 RNA was detected only in the NTS specimen and not in the gargle specimen in 3 of the NTS/ gargle pairs (Table 1). Of the 61 remaining NTS/ saliva pairs after discounting inhibited samples (6/67 for saliva), 57 of the 61 NTS/ saliva pairs were concordant (32 positive and 25 negative pairs). SARS-CoV-2 RNA was detected only in the NTS specimen in 3 of the NTS/ saliva pairs. Notably there was one saliva sample that tested positive for SARS-CoV-2 RNA while the paired NTS and gargle were negative (Table 1). Both E and S genes were detected in this positive saliva specimen (Cq values 31.46 and 31.77 respectively).

**Table 1:** Comparison of gargle vs paired NTS and saliva vs paired NTS for the detection of SARS-CoV-2 RNA.

| Gargle |  |  |  | Saliva |  |  |  |
| --- | --- | --- | --- | --- | --- | --- | --- |
|  | NTS |  |  |  | NTS |  |  |
| Gargle | Positive | Negative | Total |  | Positive | Negative | Total |
| Positive | 34 | 0 | 34 | Positive | 32 | 1 | 33 |
| Negative | 3 | 28 | 31 | Negative | 3 | 25 | 28 |
| Total | 37 | 28 | 65 | Total | 35 | 26 | 61 |
| Sensitivity | 91.9% (95% CI, 78.1% - 98.3%) |  |  | Sensitivity | 91.4% (95% CI, 76.9% - 98.2%) |  |  |
| Specificity | 100.0 % (95% CI, 87.7 % - 100.0%) |  |  | Specificity | 96.2% (95% CI, 80.4% - 99.9%) |  |  |

No significant differences were observed in the Cq values between corresponding saliva and gargle specimens (Figure 3). However, there were five discrepant saliva/gargle pairs (3 positive saliva specimens with a negative corresponding gargle specimen and 2 positive gargle specimens with a corresponding negative saliva specimen). These positive discrepant specimens all had Cq values that were within the interquartile range for positives of that sample type. Notably, there was also one saliva sample that tested positive for SARS-CoV-2 RNA while the corresponding NTS and gargle were negative.

**Figure 3.**
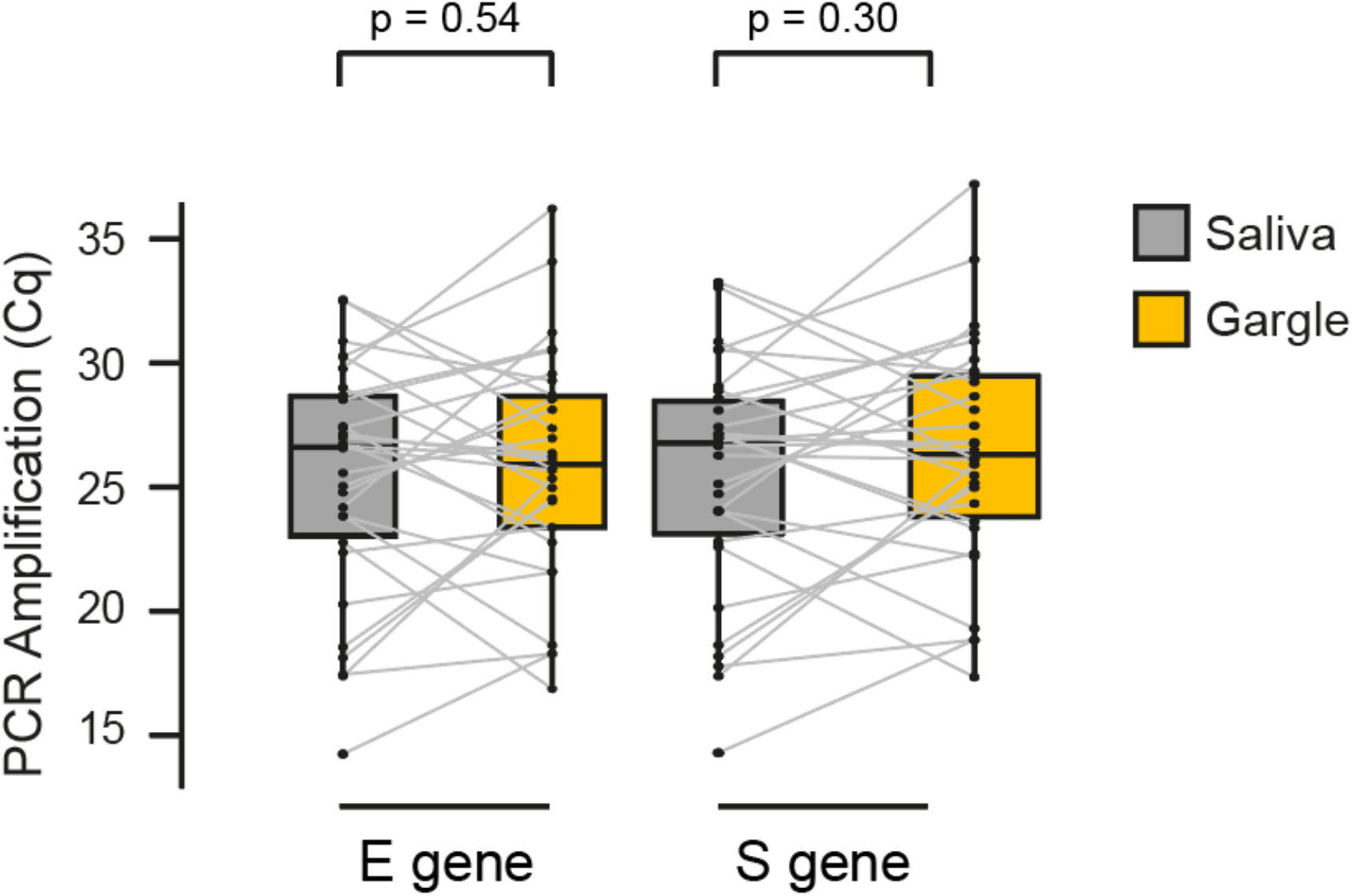
SARS-CoV-2 RNA amplification in saliva and gargle samples. Comparison between Cq values (E gene and S gene) in paired saliva and gargle samples.

Of the 261 patients who participated in Phase 2, 133 (51%) preferred the gargle method, 109 (41.7%) preferred the saliva method, and 19 (7.3%) preferred the nasopharyngeal swab method (Table 2) with no apparent gender specific differences (Supplementary Table 4). Similarly, there was no bias in sample test method according to age (Supplementary Table 5).

**Table 2.**
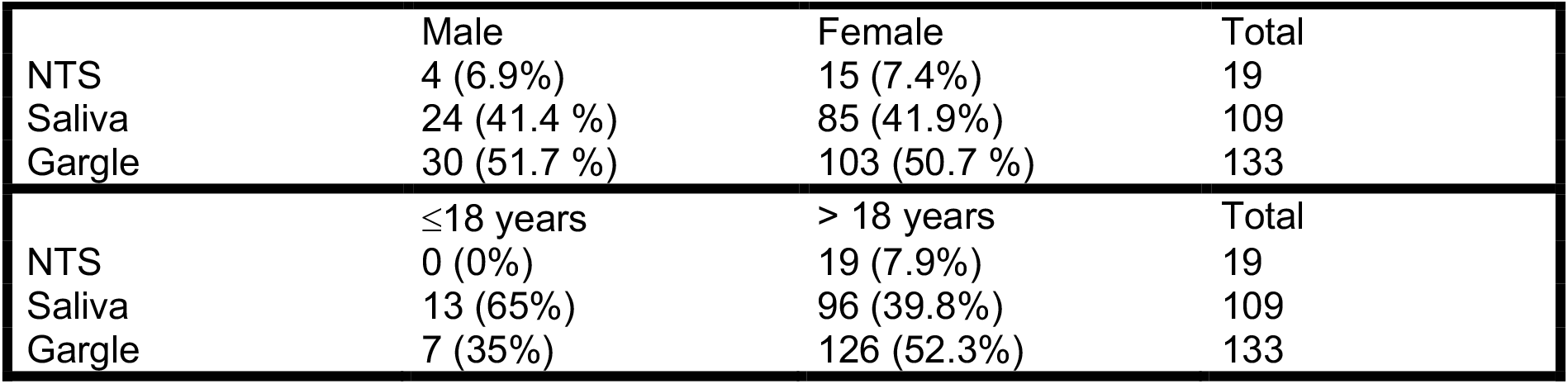
Preferential testing method stratified by gender and age.

|  | Male | Female | Total |
| --- | --- | --- | --- |
| NTS | 4 (6.9%) | 15 (7.4%) | 19 |
| Saliva | 24 (41.4 %) | 85 (41.9%) | 109 |
| Gargle | 30 (51.7 %) | 103 (50.7 %) | 133 |

|  | ≤18 years | > 18 years | Total |
| --- | --- | --- | --- |
| NTS | 0 (0%) | 19 (7.9%) | 19 |
| Saliva | 13 (65%) | 96 (39.8%) | 109 |
| Gargle | 7 (35%) | 126 (52.3%) | 133 |

## Discussion

Saliva and gargle specimens demonstrated high levels of concordance when compared with NTS specimens which corresponds well with previous studies (saliva sensitivity 93.1% (95% CI, 75.8% - 98.8%) phase 1 and 91.4% (95% CI, 76.9% - 98.2%) phase 2), gargle sensitivity 91.9% (95% CI,78.1% -98.3%)). This shows both saliva and gargle to be reliable alternative testing modalities to NTS for detection of SARS-CoV-2.

In Phase 1a a positive saliva specimen was detected where the corresponding paired NTS was negative, and similarly in Phase 2 a positive saliva specimen was detected with corresponding negative NTS and gargle specimens. Both of these positive saliva specimens had relatively high Cq values (>30 for each gene tested). Although these samples were considered as false positives, both saliva specimens could be true positive cases as despite being weakly positive all three genes were detected in the positive saliva specimen in Phase 1a, and both E and S gene detected in the positive saliva specimen in Phase 2. The potential for increased sensitivity of saliva compared to NTS has also been described previously^13^.

In Phase 1a there was a relatively high level of amplification failure for saliva (7.3%) compared to NTS (0.9%) samples. One possible explanation is the high viscosity of saliva which increases the complexity of specimen handling and requires additional pre-processing steps in the lab to overcome this issue. Since undertaking this study we, and others, have explored alternate methods for reducing saliva sample viscosity including the addition of DTT, proteinase K and sample agitation by vortexing. In contrast to saliva, gargle samples do not have the same challenges but instead produce larger volumes of fluid which could be more difficult for lab handling on automated systems, and may increase the risk of spillage. As there was no significant difference in Cq values detected between saliva samples stored and transported at 4°C vs ambient temperature, cold transport is not required which increases the practicality of these sample types.

In contrast to NTS, self-collected saliva and gargle samples are easy to obtain, and more acceptable to patients, with the distinct advantage of being a less invasive testing modality. Sampling with these methods also obviates the need for contact with a healthcare professional and reduces the use of PPE and other resources at testing centres in the face of pervasive testing supply shortages. Home self-sampling using these sample types would avoid the requirement for symptomatic individuals to attend testing facilities and reduce risk of viral transmission to others. This would have particular utility in rural settings where testing facilities are less available. Furthermore, the use of these sample types could increase compliance with testing and screening programmes, particularly those who are required to undergo regular asymptomatic screening. Their non-invasive nature may also remove some of the difficulties surrounding consent for and compliance with NTS in populations such as young children and those with cognitive impairment.

Overall gargle specimens were the most acceptable test. This was irrespective of sex with 50.7% of females and 51.7% of males choosing the gargle as their preferred sample method. Saliva was preferred by 41.9% of females and 41.8% of males, while NTS was the most acceptable in only 7.4% of females and 6.9% of males. Of those aged 18 years and under, 65% preferred saliva testing and 35% preferred gargle with none selecting NTS as their preferred testing method. Of those aged >18 years 52.3% preferred gargle testing, 39.8% preferred saliva and 7.9% preferred NTS. Using Fisher’s Exact Test, there was no significant association between gender and sample collection method or between age and sample collection method (Supplementary Tables 4 and 5).

Study participants preferring the gargle and saliva samples cited ease of performance and reduced discomfort compared with NTS as reasons for this response. Some individuals chose gargle over saliva as they felt that the saliva sample took longer to produce, whereas the gargle was quicker. Other participants found the saltiness of the saline solution unpleasant and for that reason preferred the saliva test. Those who preferred the NTS offered a variety of explanations including ease, speed, being used to it, the perception of a more accurate result, and being less unpleasant than they had expected. Of important note, the volume of saliva required for this study was greater than that which would be necessary in practice (0.5 – 1 ml), and reducing the volume required may further increase the acceptability of saliva testing.

There is limited available literature comparing the validity and acceptability of both saliva and gargle specimens with NTS. One study by Goldfarb et al[20] carried out in children aged 4-12 years found that gargle was significantly more sensitive than saliva when compared to NTS. However, the order of sample collection was alternated which may be a confounding factor as performing mouth rinse prior to saliva sampling is likely to dilute the saliva specimen and thus decrease it’s sensitivity. In our study a saliva specimen was obtained prior to saline gargle in all participants. Goldfarb et al found gargle to be more acceptable than saliva or NTS testing in their study population which is consistent with our findings.

A degree of compliance is required to provide a saliva or gargle sample and further work is required to explore the feasibility of alternative sample collection techniques in individuals unable to comply with the instructions required. Some individuals may also be unable to produce sufficient saliva including those with conditions such as sicca syndrome, or those taking medications that cause xerostomia.

## Conclusions

Our study confirms that both saliva and gargle sample types are suitable for use as an alternative testing modality to NTS, particularly in scenarios where the latter cannot be obtained, and for individuals required to undergo repeat asymptomatic screening. These samples are sufficiently stable at room temperature to allow ambient transport to the lab. The option of these alternative sampling techniques increases diagnostic capacity and versatility in the face of ongoing significant testing demands.

## Methods

### Study Phase 1a and 1b: Saliva sampling

NHS Lothian Health Care Workers (HCWs) or their symptomatic household contacts attending the drive through NHS Lothian COVID-19 testing Centre were offered a saliva test in addition to their routine NTS. Phase 1a took place between 20^th^ – 22^nd^ May 2020 at the Chalmers Hospital, Edinburgh, and Phase 1b took place between 5^th^ – 16^th^ October 2020 at the Western General Hospital, Edinburgh. In May the predominant variant was clade 20A whilst in October the predominant variant was clade 20E (EU1). These individuals had been referred for testing via the NHS Lothian Occupational Health Department. Children under the age of 5 years were excluded due to the level of compliance required to produce the specimen. Individuals were also excluded if they had eaten, had a drink, smoked, chewed gum, or brushed their teeth within the 30 minute period preceding the test. A written information leaflet was provided to each eligible attendee and verbal consent for involvement was obtained prior to participation in the study. Paired nasopharyngeal and oropharyngeal specimens (referred to as nose/ throat swabs (NTS) for the purposes of this paper) were obtained by trained testing centre staff prior to saliva testing. Those who agreed to take part were asked to produce a saliva sample by repeatedly pooling saliva in their mouth and spitting into a universal specimen container. In Phase 1a participants were asked to provide one 5 ml saliva sample; these specimens were transported to the lab by cold chain in coolboxes with ice packs. During Phase 1b participants were asked to produce 2 saliva samples (2 ml saliva per container), one stored and transported in a 4°C refrigerator, and the other at ambient temperature.

### Study Phase 2: Saliva and Gargle sampling

The second phase of the study took place between the 2^nd^ – 13^th^ November 2020. NHS Lothian HCWs or their symptomatic household contacts attending the drive through COVID testing centre at the Western General Hospital, Edinburgh were offered both saliva and pharyngeal gargle tests in addition to routine upper respiratory swab testing. Children aged 5 years or less were once again excluded along with individuals who had had eaten, had a drink, smoked, chewed gum, or brushed their teeth within the 30 minute period preceding the test. A written information leaflet was provided to each eligible attendee and verbal consent was obtained as per phase 1. NTS specimens were obtained by testing centre staff prior to saliva and gargle specimens. Participants were asked to provide one saliva sample and one gargle sample. Saliva was obtained as per phase 1 but only one 2 ml sample was required in phase 2. For gargle specimens, participants were asked to gargle 10 ml of 0.9% saline for 20 seconds then deposit the gargle liquid into a universal specimen container. The saliva and gargle specimens were transported to the lab at ambient temperature.

### User acceptability of sampling

Participants who provided all 3 specimen types in Phase 2 were asked to select their preferred testing modality and to provide reasons for their choice.

### Laboratory processing

#### Phase 1a: Saliva sampling (cold storage and transport of saliva specimens)

Saliva and corresponding NTS specimens were processed at the Institute of Genetics and Cancer (IGC) Laboratories on the Western General Hospital Campus, Edinburgh. Existing equipment and reagents were used as per previously validated protocol for COVID-19 RT-qPCR using Thermofisher TaqPath CE-IVD kits^25^. 200 μl saliva or NTS specimen was lysed with 250 μl TNA lysis buffer (Omega Biotek) containing carrier and control RNA. The saliva samples were treated with proteinase K, then each sample extracted using the Omega Biotek MAG-BIND® VIRAL DNA/RNA kit on a Thermofisher Kingfisher Flex according to the supplier’s Supplementary Protocol for NP Swabs (April 2020 version). Testing was performed using a ABI TaqPath™ COVID-19 Multiplex Assay for the N, ORF and S genes on a ABI 7500 Fast Real-Time PCR machines^25^.

#### Phase 1b: Saliva sampling (ambient/ cold storage and transport of saliva specimens)

Saliva and corresponding NTS specimens were processed at the Royal Infirmary of Edinburgh. Two shipping conditions were used to evaluate the stability of SARS-CoV-2 in saliva samples between collection and receipt in the laboratory for testing. Total nucleic acid extraction was conducted on the bioMerieux easyMAG® or EMAG® (bioMerieux Inc, Durham, NC); briefly, for all individual specimens tested, 200 μl of the sample was added to 2 ml NucliSENS Lysis Buffer (bioMerieux) and extracted into 110 μl of eluate. Testing was performed for the E and S genes on ABI 7500FAST Dx instruments using the RealStar® SARS-CoV-2 RT-PCR Kit (Altona-Diagnostics) according to the manufacturer’s instructions. Saliva samples were pre-treated with proteinase K whereby 200 μl of sample was mixed with 25 μl of molecular grade proteinase K (NEB) and then inactivated by heating at 95°C for 10 min prior to extraction.

#### Phase 2: Saliva, and Gargle sampling

Saliva samples were processed as per Phase 1b. Gargle samples (1 ml) were mixed with 1 ml VPSS (Viral PCR Sample Solution, E&O Laboratories; 53% guanidine thiocyanate, 44 mM Tris-HCl pH 6.4, 20 mM EDTA, 25 TX-100) and incubated for 10 min to ensure inactivation of virus before proceeding to extraction as described above.

Discrepant samples were tested for the *RPP30* gene, which encodes the human RNase P protein subunit P30[25].

## Statistical Analysis

The diagnostic accuracy of saliva and gargle samples was determined by estimating sensitivity and specificity with exact binomial 95% confidence intervals (CIs) using detection rate in NTS as the gold standard. The significance of sample type/shipping conditions on Cq values was determined using the Wilcoxon Test for paired samples and the results plotted using the ggpubr package (v.0.4.0) for R. All analyses were performed using R software (ver. 4.0.3). The effect of gender and age on sample collection method choice was assessed using Fisher’s Exact Test.

## Data Availability

All data produced in the present work are contained in the manuscript

## Acknowledgements

The authors would like to thank the clinical staff at the Chalmers Hospital/ Western General Hospital COVID-19 Testing Centres who collected samples from patients and the laboratory staff at the MRC Institute of Genetics and Cancer and the Royal Infirmary of Edinburgh who processed samples. This work was part of the NHS Laboratories Programme COVID-19 innovation saliva testing short-life working group with representation from the NHS Lothian Occupational Health and Safety Service, NHS Lothian Laboratories, the University of Edinburgh, NHS National Services Scotland, and the Scottish Government, as well as Clinical Scientists and representatives from NHS Lothian, NHS Greater Glasgow and Clyde, NHS Grampian, NHS Lanarkshire, and NHS Fife. The authors would like to acknowledge the help and support provided by all members of this group.

---

**Supplementary Table 1.** Full results for Phase 1a NTS and Saliva samples showing amplification (Cq). U = Undetermined. Green, positive NTS samples; beige, failed samples; blue, inconclusive samples

| Specimen Number | TaqPath NTS | Amplification (Cq) |  |  |  | TaqPath Saliva | Amplification (Cq) |  |  |  |
| --- | --- | --- | --- | --- | --- | --- | --- | --- | --- | --- |
|  | Result | MS2 | Ngene | ORFgene | Sgene | Result | MS2 | Ngene | ORF | Sgene |
| 1 | Negative | 23.64 | U | U | U | Negative | 24.606 | U | U | U |
| 2 | Negative | 24.474 | U | U | U | Fail | U | U | U | U |
| 3 | Negative | 24.34 | U | U | U | Negative | 24.948 | U | U | U |
| 4 | Negative | 24.149 | U | U | U | Negative | 24.534 | U | U | U |
| 5 | Positive | 24.855 | 24.145 | 23.366 | 24.422 | Positive | 25.025 | 26.834 | 26.242 | 26.67 |
| 6 | Negative | 25.099 | U | U | U | Negative | 24.215 | U | U | U |
| 7 | Negative | 25.015 | U | U | U | Fail | U | U | U | U |
| 8 | Negative | 24.745 | U | U | U | Negative | 24.023 | U | U | U |
| 9 | Negative | 25.485 | U | U | U | Negative | 25.103 | U | U | U |
| 10 | Negative | 24.606 | U | U | U | Negative | 25.859 | U | U | U |
| 11 | Negative | 25.376 | U | U | U | Negative | 23.628 | U | U | U |
| 12 | Positive | 24.715 | 24.023 | 24.951 | 25.551 | Positive | 24.569 | 20.562 | 20.922 | 21.985 |
| 13 | Negative | 24.298 | U | U | U | Negative | 24.077 | U | U | U |
| 14 | Negative | 24.612 | U | U | U | Negative | 24.975 | U | U | U |
| 15 | Positive | 24.471 | 28.526 | 27.939 | 28.234 | Positive | 25.527 | 33.859 | 32.922 | 33.688 |
| 16 | Negative | 24.253 | U | U | U | Negative | 25.801 | U | U | U |
| 17 | Negative | 24.774 | U | U | U | Negative | 26.002 | U | U | U |
| 18 | Negative | 24.514 | U | U | U | Negative | 25.181 | U | U | U |
| 19 | Negative | 25.001 | U | U | U | Negative | 25.388 | U | U | U |
| 20 | Negative | 24.535 | U | U | U | Negative | 23.448 | U | U | U |
| 21 | Negative | 24.201 | U | U | U | Negative | 25.874 | U | U | U |
| 22 | Negative | 24.368 | U | U | U | Negative | 27.561 | U | U | U |
| 23 | Negative | 24.196 | U | U | U | Negative | 24.444 | U | U | U |
| 24 | Negative | 25.153 | U | U | U | Negative | 25.565 | U | U | U |
| 25 | Negative | 24.52 | U | U | U | Negative | 24.933 | U | U | U |
| 26 | Negative | 25.344 | U | U | U | Negative | 24.808 | U | U | U |
| 27 | Negative | 24.628 | U | U | U | Negative | 26.142 | U | U | U |
| 28 | Negative | 23.868 | U | U | U | Negative | 26.017 | U | U | U |
| 29 | Negative | 23.806 | U | U | U | Negative | 25.125 | U | U | U |
| 30 | Negative | 23.946 | U | U | U | Negative | 24.39 | U | U | U |
| 31 | Negative | 24.269 | U | U | U | Negative | 24.372 | U | U | U |
| 32 | Negative | 24.53 | U | U | U | Negative | 24.197 | U | U | U |
| 33 | Negative | 24.852 | U | U | U | Negative | 24.54 | U | U | U |
| 34 | Negative | 25.703 | U | U | U | Negative | 25.323 | U | U | U |
| 35 | Negative | 26.76 | U | U | U | Fail | 35.686 | U | U | U |
| 36 | Negative | 26.005 | U | U | U | Negative | 24.164 | U | U | U |
| 37 | Negative | 25.478 | U | U | U | Negative | 24.6 | U | U | U |
| 38 | Negative | 25.569 | U | U | U | Negative | 23.101 | U | U | U |
| 39 | Negative | 25.791 | U | U | U | Negative | 24.134 | U | U | U |
| 40 | Negative | 24.741 | U | U | U | Negative | 24.142 | U | U | U |
| 41 | Negative | 25.468 | U | U | U | Negative | 24.935 | U | U | U |
| 42 | Negative | 24.729 | U | U | U | Inconclusive | 22.603 | 35.974 | U | U |
| 43 | Negative | 25.233 | U | U | U | Negative | 25.125 | U | U | U |
| 44 | Negative | 26.097 | U | U | U | Negative | 25.231 | U | U | U |
| 45 | Negative | 24.328 | U | U | U | Negative | 25.942 | U | U | U |
| 46 | Negative | 25.652 | U | U | U | Negative | 24.277 | U | U | U |
| 47 | Negative | 27.159 | U | U | U | Negative | 24.433 | U | U | U |
| 48 | Negative | 25.397 | U | U | U | Negative | 25.033 | U | U | U |
| 49 | Negative | 25.334 | U | U | U | Negative | 24.843 | U | U | U |
| 50 | Negative | 24.726 | U | U | U | Negative | 24.731 | U | U | U |
| 51 | Negative | 24.387 | U | U | U | Negative | 25.505 | U | U | U |
| 52 | Negative | 24.402 | U | U | U | Negative | 25.129 | U | U | U |
| 53 | Negative | 24.729 | U | U | U | Negative | 23.928 | U | U | U |
| 54 | Negative | 25 | U | U | U | Negative | 25.269 | U | U | U |
| 55 | Negative | 24.445 | U | U | U | Negative | 23.924 | U | U | U |
| 56 | Negative | 24.699 | U | U | U | Negative | 24.324 | U | U | U |
| 57 | Negative | 25.35 | U | U | U | Negative | 25.254 | U | U | U |
| 58 | Positive | 25.007 | 25.095 | 25.764 | 25.924 | Positive | 23.96 | 26.168 | 26.156 | 27.145 |
| 59 | Negative | 24.899 | U | U | U | Negative | 23.146 | U | U | U |
| 60 | Negative | 25.357 | U | U | U | Negative | 24.243 | U | U | U |
| 61 | Negative | 26.072 | U | U | U | Fail | 34.529 | U | U | U |
| 62 | Fail | 31.002 | U | U | U | Negative | 24.667 | U | U | U |
| 63 | Negative | 24.707 | U | U | U | Negative | 23.902 | U | U | U |
| 64 | Negative | 24.565 | U | U | U | Negative | 24.889 | U | U | U |
| 65 | Positive | 24.289 | 20.528 | 20.928 | 21.099 | Positive | 23.775 | 27.193 | 26.336 | 26.803 |
| 66 | Negative | 26.113 | U | U | U | Negative | 25.473 | U | U | U |
| 67 | Negative | 24.451 | U | U | U | Negative | 25.251 | U | U | U |
| 68 | Negative | 25.1 | U | U | U | Negative | 25.565 | U | U | U |
| 69 | Negative | 24.54 | U | U | U | Negative | 27.163 | U | U | U |
| 70 | Negative | 25.235 | U | U | U | Negative | 22.892 | U | U | U |
| 71 | Negative | 24.475 | U | U | U | Negative | 23.75 | U | U | U |
| 72 | Negative | 24.335 | U | U | U | Negative | 24.685 | U | U | U |
| 73 | Negative | 24.895 | U | U | U | Fail | 35.5 | U | U | U |
| 74 | Negative | 24.979 | U | U | U | Negative | 24.719 | U | U | U |
| 75 | Negative | 25.409 | U | U | U | Negative | 23.793 | U | U | U |
| 76 | Negative | 25.154 | U | U | U | Negative | 24.986 | U | U | U |
| 77 | Negative | 25.358 | U | U | U | Negative | 24.718 | U | U | U |
| 78 | Negative | 24.502 | U | U | U | Positive | 23.545 | 33.576 | 34.473 | 37.553 |
| 79 | Negative | 24.42 | U | U | U | Negative | 25.14 | U | U | U |
| 80 | Negative | 23.689 | U | U | U | Negative | 23.348 | U | U | U |
| 81 | Negative | 25.347 | U | U | U | Negative | 25.147 | U | U | U |
| 82 | Negative | 24.663 | U | U | U | Negative | 24.586 | U | U | U |
| 83 | Negative | 25.873 | U | U | U | Negative | 25.081 | U | U | U |
| 84 | Negative | 24.905 | U | U | U | Negative | 25.593 | U | U | U |
| 85 | Positive | 24.793 | 23.678 | 23.384 | 24.585 | Positive | 24.865 | 22.524 | 21.686 | 22.499 |
| 86 | Negative | 23.678 | U | U | U | Negative | 25.117 | U | U | U |
| 87 | Negative | 24.283 | U | U | U | Negative | 24.928 | U | U | U |
| 88 | Negative | 24.832 | U | U | U | Fail | 28.495 | U | U | U |
| 89 | Negative | 24.159 | U | U | U | Negative | 24.702 | U | U | U |
| 90 | Negative | 24.228 | U | U | U | Negative | 25.553 | U | U | U |
| 91 | Negative | 24.464 | U | U | U | Fail | 34.225 | U | U | U |
| 92 | Positive | 24.969 | 35.225 | 36.634 | 38.617 | Positive | 30.036 | 39.775 | 35.627 | U |
| 93 | Positive | 24.643 | 23.633 | 24.407 | 24.711 | Positive | 25.238 | 24.068 | 23.744 | 24.5 |
| 94 | Positive | 25.635 | 30.696 | 29.768 | 29.619 | Positive | 23.179 | 22.117 | 21.421 | 22.124 |
| 95 | Negative | 25.605 | U | U | U | Negative | 26.102 | U | U | U |
| 96 | Negative | 25.337 | U | U | U | Negative | 23.301 | U | U | U |
| 97 | Negative | 24.281 | U | U | U | Negative | 26.012 | U | U | U |
| 98 | Negative | 24.932 | U | U | U | Negative | 24.823 | U | U | U |
| 99 | Negative | 24.67 | U | U | U | Negative | 24.58 | U | U | U |
| 100 | Negative | 25.797 | U | U | U | Fail | 27.764 | U | U | U |
| 101 | Negative | 24.457 | U | U | U | Negative | 24.8 | U | U | U |
| 102 | Negative | 25.254 | U | U | U | Negative | 26.826 | U | U | U |
| 103 | Negative | 24.284 | U | U | U | Negative | 24.68 | U | U | U |
| 104 | Negative | 24.581 | U | U | U | Negative | 24.917 | U | U | U |
| 105 | Negative | 25.411 | U | U | U | Negative | 26.762 | U | U | U |
| 106 | Negative | 24.642 | U | U | U | Negative | 25.418 | U | U | U |
| 107 | Negative | 26.032 | U | U | U | Negative | 24.445 | U | U | U |
| 108 | Positive | 23.944 | 26.894 | 26.625 | 26.808 | Positive | 23.821 | 26.122 | 25.271 | 26.434 |
| 109 | Negative | 24.718 | U | U | U | Negative | 25.631 | U | U | U |
|  | average | 24.925 |  |  |  | average | 25.297 |  |  |  |
|  | stdev | 0.8651 |  |  |  | stdev | 2.2137 |  |  |  |
|  | average-2SD | 23.194 |  |  |  | average-2SD | 20.87 |  |  |  |
|  | average+2SD | 26.655 |  |  |  | average+2SD | 29.724 |  |  |  |
|  | minimum | 23.64 |  |  |  | minimum | 22.603 |  |  |  |

**Supplementary Table 2.** Sample results from Phase 1b saliva and paired NTS testing (NTS = nose throat swab, IC = internal control). Green, NTS positive; beige, discordant result.

| Sample Number | Ambient |  |  |  | Cold Chain |  |  |  | NTS Result |
| --- | --- | --- | --- | --- | --- | --- | --- | --- | --- |
|  | E gene | S gene | IC | Result | E gene | S gene | IC | Result |  |
| MI611007W | 26.6 | 27.3 | 29.9 | Positive | 24 | 24.5 | 32.9 | Positive | Positive |
| MI611104S | 29.2 | 30.3 | 31 | Positive | 28.2 | 29.1 | 31.2 | Positive | Positive |
| MI611105G | 25.2 | 25.7 | - | Positive | 26.6 | 26.7 | - | Positive | Positive |
| MI611117G | 18.4 | 19 | 34.2 | Positive | 26.8 | 27.2 | 33.2 | Positive | Positive |
| MI611078S | 25 | 25.5 | - | Positive | 14.4 | 14.5 | - | Positive | Positive |
| MI611119N | 33 | 35.9 | 29.5 | Positive | 31.7 | - | 32 | Positive<sup>§</sup> | Positive |
| MI611120R | 23.6 | 24.2 | 31.5 | Positive | 23.9 | 24.5 | 31.3 | Positive | Positive |
| MI611132X | 25.4 | 26.1 | 35 | Positive | 24.9 | 25.4 | - | Positive | Positive |
| MI611147S | 20.8 | 21.3 | 31.5 | Positive | 16 | 16.3 | 37.8 | Positive | Positive |
| MI611159S | 29.9 | 30.8 | 30.3 | Positive | 29.2 | 29.8 | 31.8 | Positive | Positive |
| MI611162K | 32.9 | - | 29.5 | Positive<sup>§</sup> | 35.8 | - | 30 | Positive<sup>§</sup> | Positive |
| MI611168X | 22.3 | 23.1 | 33.7 | Positive | 21.9 | 22.6 | 36.5 | Positive | Positive |
| MI611175M | - | - | 30.5 | Negative | - | - | 31.1 | Negative | Positive |
| MI611185T | 24.8 | 25.4 | 31.7 | Positive | 26.3 | 27.1 | 30.9 | Positive | Positive |
| MI611189V | 26.7 | 27.3 | - | Positive | 28.1 | 28.8 | 34 | Positive | Positive |
| MI611196C | 26.7 | 27.2 | 34.5 | Positive | 28.1 | 28.8 | 29.7 | Positive | Positive |
| MI611202Z | 28.5 | 29.1 | 31.1 | Positive | 30.3 | 31.2 | 30 | Positive | Positive |
| MI611219Y | 29.6 | 30 | 29.1 | Positive | 33 | 33.3 | 28.6 | Positive | Positive |
| MI611224E | - | - | 29.8 | Negative | - | - | 30.2 | Negative | Positive |
| MI611124G | - | - | 29.8 | Negative | - | - | 30.2 | Negative | Negative |
| MI611123S | - | - | 29.1 | Negative | - | - | 29 | Negative | Negative |
| MI611122L | - | - | 29.5 | Negative | - | - | 29.2 | Negative | Negative |
| MI611121X | - | - | 29.8 | Negative | - | - | 29.4 | Negative | Negative |
| MI611118Z | - | - | 30 | Negative | - | - | 31.4 | Negative | Negative |
| MI611116S | - | - | 31.3 | Negative | - | - | 32.4 | Negative | Negative |

<sup>§</sup>Confirmed as positive by testing with the CDC N2 gene assay

**Supplementary Table 3.** Full results from Phase 2 gargle, saliva and dependent NTS testing (NTS = nose throat swab, IC = internal control). Green, NTS positive; beige, discordant result.

| Sample Number | Saliva |  |  |  | Gargle |  |  |  | NTS Result |
| --- | --- | --- | --- | --- | --- | --- | --- | --- | --- |
|  | E gene | S gene | IC | Result | E gene | S gene | IC | Result |  |
| MI611226Y | - | - | 29.7 | Negative | - | - | 29.29 | Negative | Negative |
| MI611228P | - | - | 29.22 | Negative | - | - | 29.06 | Negative | Negative |
| MI611229A | - | - | 30.55 | Negative | - | - | 29.56 | Negative | Negative |
| MI611230D | - | - | 29.13 | Negative | - | - | 29.37 | Negative | Negative |
| MI611231L | - | - | 28.94 | Negative | - | - | 28.51 | Negative | Negative |
| MI611232G | - | - | 30.03 | Negative | - | - | 29.48 | Negative | Negative* |
| MI611233Z | - | - | - | Inhibited | - | - | 29.04 | Negative | Negative |
| MI611235Q | - | - | - | Inhibited | - | - | 31.48 | Negative | Negative |
| MI611237H | - | - | 29.27 | Negative | - | - | 30.65 | Negative | Negative |
| MI611239W | - | - | - | Inhibited | - | - | 30.72 | Negative | Negative |
| MI611240R | - | - | 28.05 | Negative | - | - | 31.03 | Negative | Negative |
| MI611243S | 31.46 | 31.77 | 28.18 | Positive | - | - | 30.94 | Negative | Negative |
| MI611244G | - | - | 30.38 | Negative | - | - | 30.53 | Negative | Negative |
| MI611245Z | - | - | - | Inhibited | - | - | 31.01 | Negative | Negative |
| MI611421K | - | - | 27.96 | Negative | - | - | 29.13 | Negative | Negative |
| MI611422J | - | - | 27.69 | Negative | - | - | 29.67 | Negative | Negative |
| MI611423V | - | - | 29.43 | Negative | - | - | 30.07 | Negative | Negative |
| MI611424B | - | - | 28.34 | Negative | - | - | 33.03 | Negative | Negative |
| MI611425R | - | - | 30.27 | Negative | - | - | 29.31 | Negative | Negative |
| MI611426X | - | - | 27.99 | Negative | - | - | - | Inhibited | Negative |
| MI611427D | - | - | 29.15 | Negative | - | - | 29.38 | Negative | Negative |
| MI611430F | - | - | 33.07 | Negative | - | - | 30.02 | Negative | Negative |
| MI611480G | - | - | 28.17 | Negative | - | - | 29.53 | Negative | Negative |
| MI611481Z | - | - | 27.77 | Negative | - | - | 29.56 | Negative | Negative |
| MI611482Q | - | - | 27.96 | Negative | - | - | 28.86 | Negative | Negative |
| MI611483E | - | - | 28.4 | Negative | - | - | 29.31 | Negative | Negative |
| MI611484H | - | - | 27.78 | Negative | - | - | 33.31 | Negative | Negative |
| MI611486W | - | - | 28.68 | Negative | - | - | - | Inhibited | Negative |
| MI611487P | - | - | 27.35 | Negative | - | - | 30.25 | Negative | Negative |
| MI611488A | - | - | 29.16 | Negative | - | - | 29.44 | Negative | Negative |
| MI611227W | 28.56 | 28.62 | 29.34 | Positive | 30.53 | 30.92 | 28.89 | Positive | Positive |
| MI611234N | 32.6 | 33.1 | - | Positive | 27.41 | 28.15 | 37.67 | Positive | Positive |
| MI611242L | 14.26 | 14.31 | - | Positive | 18.31 | 18.86 | 34.09 | Positive | Positive |
| MI611246N | 25.6 | 26.9 | - | Positive | 27 | 27.51 | 31.1 | Positive | Positive |
| MI611254L | 18.15 | 18.2 | - | Positive | 24.57 | 25.19 | 30.75 | Positive | Positive |
| MI611260M | 23.86 | 24.06 | - | Positive | 21.62 | 22.24 | 33.27 | Positive | Positive |
| MI611261J | 17.47 | 17.8 | - | Positive | 18.3 | 18.86 | 34.99 | Positive | Positive |
| MI611263R | 27.04 | 26.7 | - | Positive | 26.4 | 26.84 | 31.53 | Positive | Positive |
| MI611265D | 30.3 | 30.9 | - | Positive | 25.75 | 26.18 | 31.01 | Positive | Positive |
| MI611267S | 26.68 | 27 | - | Positive | 26.26 | 26.79 | 31.16 | Positive | Positive |
| MI611268G | - | - | 30.76 | Negative | 23.92 | 24.4 | 30.56 | Positive | Positive |
| MI611272J | 20.3 | 20.16 | 31.83 | Positive | 21.59 | 22.35 | 38.44 | Positive | Positive |
| MI611277D | - | - | 28.33 | Negative | - | - | 30.57 | Negative | Positive |
| MI611302X | 18.57 | 18.65 | 30.12 | Positive | 24.47 | 25.01 | 31.57 | Positive | Positive |
| MI611304L | 24.21 | 24.09 | 30.95 | Positive | 31.26 | 31.54 | 32.67 | Positive | Positive |
| MI611315D | 29.82 | 29.09 | 29.4 | Positive | 36.26 | 37.26 | 30.37 | Positive | Positive |
| MI611319Z | 30.91 | 30.56 | 29.08 | Positive | 29.32 | 29.72 | 30.48 | Positive | Positive |
| MI611328Q | 28.74 | 28.13 | 29.93 | Positive | 30.61 | 31.24 | 31.23 | Positive | Positive |
| MI611339N | 25.07 | 25.16 | 29.26 | Positive | 28.15 | 28.69 | 30.36 | Positive | Positive |
| MI611342V | 27.42 | 27.46 | 34.39 | Positive | 29.59 | 29.6 | 30.59 | Positive | Positive |
| MI611345X | 22.39 | 22.83 | 30.17 | Positive | 23.41 | 24.37 | 30.95 | Positive | Positive |
| MI611354V | 22.8 | 22.62 | 29.56 | Positive | 16.89 | 17.36 | 36.02 | Positive | Positive |
| MI611356R | 26.6 | 26.31 | 30.32 | Positive | 25.7 | 26.15 | 30.63 | Positive | Positive |
| MI611358D | 26.79 | 27.1 | 28.19 | Positive | 23.4 | 23.65 | 31.85 | Positive | Positive |
| MI611366V | - | - | - | Inhibited | 34.12 | 34.62 | 30.86 | Positive | Positive |
| MI611367B | 27.16 | 27.18 | 30.07 | Positive | 26.13 | 26.5 | 30.8 | Positive | Positive |
| MI611380H | 23.26 | 23.28 | 28.83 | Positive | - | - | 29.24 | Negative | Positive |
| MI611383A | 28.19 | 28.14 | 36.79 | Positive | - | - | 29.38 | Negative | Positive |
| MI611401T | 30.3 | 30.6 | - | Positive | 34.12 | 34.21 | 29.28 | Positive | Positive |
| MI611406R | 27.48 | 27.15 | 28.8 | Positive | 22.8 | 23.38 | 30.35 | Positive | Positive |
| MI611407X | - | - | - | Inhibited | 25.81 | 26.32 | 29.31 | Positive | Positive |
| MI611408D | 32.54 | 33.3 | 28.47 | Positive | 28.74 | 29.27 | 31.32 | Positive | Positive |
| MI611411F | 29.04 | 28.93 | 29.24 | Positive | 25 | 25.5 | 29.71 | Positive | Positive |
| MI611414M | - | - | 30.48 | Negative | 21.37 | 22.07 | 30.69 | Positive | Positive |
| MI611479T | 17.4 | 17.4 | 34.17 | Positive | 25.38 | 25.94 | - | Positive | Positive |
| MI611485Y | 24.82 | 24.76 | 34.12 | Positive | 28.57 | 30.18 | - | Positive | Positive |
| MI611496H | 23.86 | 24.05 | 30.21 | Positive | 18.65 | 19.33 | - | Positive | Positive |

\*Sample Lost - follow-up sample negative

**Supplementary Table 4.** Association between gender and sample collection preference

|  | Male | Female | Fishers exact test |
| --- | --- | --- | --- |
| NTS | 4 | 15 | P = 1 |
| Saliva | 24 | 85 |  |
| NTS | 4 | 15 | P = 1 |
| Gargle | 30 | 103 |  |
| Saliva | 24 | 85 | P = 1 |
| Gargle | 30 | 103 |  |

**Supplementary Table 5.** Association between age and sample collection preference

|  | 18 and under | Over 18 | Fishers exact test |
| --- | --- | --- | --- |
| NTS | 0 | 19 | P = 0.2137 |
| Saliva | 13 | 96 |  |
| NTS | 0 | 19 | P = 0.597 |
| Gargle | 7 | 126 |  |
| Saliva | 13 | 96 | P = 0.0984 |
| Gargle | 7 | 126 |  |

